# Epidemiology of the early COVID-19 epidemic in Orange County, California: comparison of predictors of test positivity, mortality, and seropositivity

**DOI:** 10.1101/2021.01.13.21249507

**Authors:** Daniel M. Parker, Tim Bruckner, Verónica M. Vieira, Catalina Medina, Vladimir N. Minin, Philip L. Felgner, Alissa Dratch, Matthew Zahn, Scott M. Bartell, Bernadette Boden-Albala

**Affiliations:** University of California, Irvine, California, U.S.A.; Orange County Health Care Agency, Santa Ana, California, U.S.A.

## Abstract

COVID-19 is one of the largest public health emergencies in modern history. Here we present a detailed analysis from a large population center in Southern California (Orange County, population of 3.2 million) to understand heterogeneity in risks of infection, test positivity, and death. We used a combination of datasets, including a population-representative seroprevalence survey, to assess the true burden of disease as well as COVID-19 testing intensity, test positivity, and mortality. In the first month of the local epidemic, case incidence clustered in high income areas. This pattern quickly shifted, with cases next clustering in much higher rates in the north-central area which has a lower socio-economic status. Since April, a concentration of reported cases, test positivity, testing intensity, and seropositivity in a north-central area persisted. At the individual level, several factors (e.g., age, race/ethnicity, zip codes with low educational attainment) strongly affected risk of seropositivity and death.

## INTRODUCTION

In late 2019 an epidemic of respiratory disease (coronavirus disease 19 or COVID-19), caused by a novel coronavirus (SARS-CoV-2) emerged in Wuhan, China, and rapidly spread worldwide. COVID-19 has manifested in different ways across social, economic, and demographic groups, with regard to apparent risk of infection, disease severity, and mortality (1– 3). The elderly and those with co-morbidities are at the highest risk of severe disease (4). Many hospitalized patients require supplemental oxygen or ventilators (5), and there is a high mortality rate among those who are hospitalized (6). In many places healthcare facilities have been overwhelmed by a surge in cases and have fallen short of needed ventilators and ICU beds, resulting in massive morbidity and mortality(7,8). Availability of tests and operational barriers were limiting factors for diagnosis in parts of the U.S. during the early months of the pandemic (9).

California is the most populous state in the U.S., with an estimated 39.5 million inhabitants. Orange County (OC) is a coastal county in California, and the sixth most populous county in the country, with an estimated 3.2 million inhabitants). The first confirmed case in California (3^rd^ in the U.S.) was reported from OC. On January 31 the WHO declared a Global Health Emergency, and on February 3^rd^ the U.S. declared a public health emergency. Subsequent cases were reported in California in February, mostly among travelers. On February 26^th^, local (‘community’) transmission was first confirmed in the United States in northern California. OC had only 2 reported cases in February. By March 11 another 6 cases had been reported in OC. On March 12 there were 8 reported cases; followed by 23 on March 13; 11 each day on March 14 and 15; and 32 on March 16. This surge in cases in mid-March in OC and other counties in California triggered emergency orders by the Governor and the County Health Officer at the Orange County Health Care Agency, prohibiting public or private gatherings and also leading to school and business closures (10). Though many businesses were closed at this time, the mandated social distancing measures had exceptions in place for individuals working in “essential jobs,” which was broadly defined and included medical professionals, food providers, delivery agencies, public officials, contractors, and building laborers (11). The social and economic characteristics of individuals working essential jobs likely differs from the overall population.

Almost half of OC residents over the age of five speak a language other than English at home. Additionally, many within the Hispanic/Latino and Asian communities of OC live below the poverty level (17.9% and 16%, respectively) and face challenges in education, household income, access to healthcare, health disparities, and life expectancy (12,13). The relatively small land area, high population density, and diverse population of OC provides a unique opportunity to explore potentially important social, economic, and demographic correlates of COVID-19 epidemiology.

Here we present results from a detailed spatiotemporal epidemiological analysis of COVID-19 in OC, California. First, we draw from reported tests and mortality from the county health agency. Given that passively detected cases are prone toward bias, in July we also conducted a seroprevalence survey to assess the true burden of disease in the county. In our analyses we leverage both datasets to compare predictors of test positivity, mortality, and seropositivity over the first 6 months of the epidemic.

## METHODS

### Data

#### Case and Mortality data

Case data were provided through a memorandum of understanding with the Orange County Health Care Agency (OCHCA) and consisted of individual level records of all negative and positive PCR tests conducted throughout the county from March 1 through August 16, 2020 (this date aligns with our cross-sectional seroprevalence survey which completed on August 16). OCHCA receives testing data from the California Reportable Disease Information Exchange (CalREDIE), an infectious disease surveillance system implemented by the California Department of Public Health (CDPH) [14]. The data include information on test date, age, gender, race, ethnicity, and zip code of the individual taking the test. For individuals who had repeat PCR testing after testing positive, only the first positive diagnosis was included in our analyses. Mortality data were also provided by OCHCA and consisted of individual-level records of deaths attributed to COVID-19.

#### Seroprevalence data

Participants for the serological survey were recruited using a proprietary database maintained by SoapBoxSample, an LRW Group Company. The database is intended to be representative of the age, income, and racial/ethnic diversity of OC. Participants were contacted by email or phone. We recruited one participant per household to participate in a survey on their thoughts and opinions regarding COVID-19. The survey included questions on socio-demographics, occupation, social activities, any illness or symptoms in the last few months, and whether the individual had been diagnosed with COVID-19. After completing this portion of the survey, each eligible participant was asked if they would be willing to participate in a drive-thru blood test for SARS-CoV-2 antibodies. Eligibility for antibody testing was restricted to a quota sample designed to be demographically representative of the county as a whole. Recruitment to the antibody test was delayed to the end of the questionnaire in order to avoid biasing the serological survey toward individuals who believe that they were infected with SARS-CoV-2. There were a total of 10 field sites for drive-through blood tests, dispersed throughout OC in order to minimize driving distances for participants. The seroprevalence study design and overall findings for OC are described in more detail elsewhere (14).

#### Serological test

We used a coronavirus antigen microarray to classify participants from the serological survey as seropositive or seronegative. The array tests for both IgG and IgM and contains 12 antigens from SARS-CoV-2 (described in detail at (15)).

#### Zip-code level socio-demographic data

At the zip-code level we included median household income, the percentage of adults above age 25 who have at least a bachelor’ s degree, and the percentage of adults who have had insurance in the previous 5 years. These data come from the 2018 American Community Survey of 2018 (extracted from (12)).

### Analysis

#### Descriptive Spatiotemporal Data Analysis

Reported cases and number of tests were aggregated at the zip code level and by week. A total of 85 zip codes were included in the analysis. For plotting cases on OC maps, the data were further aggregated by months (March through August). Case incidence was calculated and mapped as positive cases per 100,000 population per week. Testing intensity was calculated and mapped as total number of tests per 100,000 people per week. Test positivity was calculated and mapped as the percentage of positive tests for each month.

Formal testing of spatial autocorrelation was done using the global Moran’ s *I* statistic and spatial correlograms. Both identify the presence and extent of clustering or dispersion. Local clustering statistics (Local Indicators of Spatial Autocorrelation or LISA (16)) were then used to visualize the location of clusters. All tests were run for case incidence, test positivity, and test intensity. Seropositivity was also mapped and assessed using LISA statistics (since this was a cross sectional survey there is no time component).

#### Relational Analysis of COVID-19 Test Positivity, Mortality, and Seropositivity

We used logistic regressions to explore geographic, demographic, economic, and epidemiological predictors of the odds of testing positive for COVID-19, of dying from COVID-19, and of being seropositive for SARS-CoV-2 antibodies. Predictors in our models are listed in **Supplementary Table 1** and included: age group, gender, and race/ethnicity at the individual level. Zip code level predictors included: median household income, the percentage of adults over age 25 with at least a bachelor’ s degree, the percentage of adults who have had insurance in the previous 5 years, and population density (individuals per square kilometer).

Several specifications were tested for model fit, interpretability, and parsimony. Through preliminary exploratory analyses we noted that the first cases were reported from coastal zip-codes but that this pattern had shifted inland over time. Given the changing dynamics over time, we explored different specifications for ‘time’ in the model for test positivity. The best fitting model included a smoothed interaction term for time (coded by day, **Supplemental Table 1**) and median household income at the zip code level.

The same predictors were included in the model for mortality, save for the interaction between time and median household income (which did not improve model performance). Given reports of increased mortality related to hospital bed shortages, we also included as a predictor the number of ICU beds occupied by suspected or confirmed COVID-19 patients on the day that an individual tested positive for SARS-CoV-2.

We compared three seropositivity models, with various combinations of individual and zip-code level predictors (see details in **Supplementary Appendix A**). All model results are presented as model adjusted odds ratios (AORs) with 95% confidence intervals (CIs). Model summary statistics, including model Bayesian Information Criterion are presented in **Supplementary Appendix A**.

### Software

Maps were created using QGIS version 3.4.9. Tests for spatial autocorrelation were done using GeoDa version 1.14.0. All other analyses were conducted using R statistical software version 3.5.2.

## RESULTS

A total of 597,922 tests were reported to OCHCA up to August 16, 2020. After dropping repeated tests and those with incomplete data, 318,492 individuals were included. Of these individuals, 36,816 tested positive for COVID-19 and 1,248 died from the disease. In the separate population-based serological survey, 2,979 individuals participated and 350 tested seropositive.

### Spatial patterns in reported COVID-19 cases, testing intensity, and seropositivity

The global Moran’ s *I* statistics and spatial correlograms indicated significant clustering in reported cases and testing intensity in the first month (March) of the local epidemic (**Table 1; Supplementary Figures 3 and 4**). Conversely, there was no detectable clustering of test positivity in March (**Table 1, Supplementary Figure 5**). The highest reported case incidence in March was along the central coast and southern portion of the county (**Figure 1 A**). The LISA statistics indicated statistically significant clustering of high incidence zip codes in the central coast area (**Figure 1 B**). This clustering of case incidence overlaps with clustering of test intensity in March (**Figure 2 A and B**).

**Table 1.**
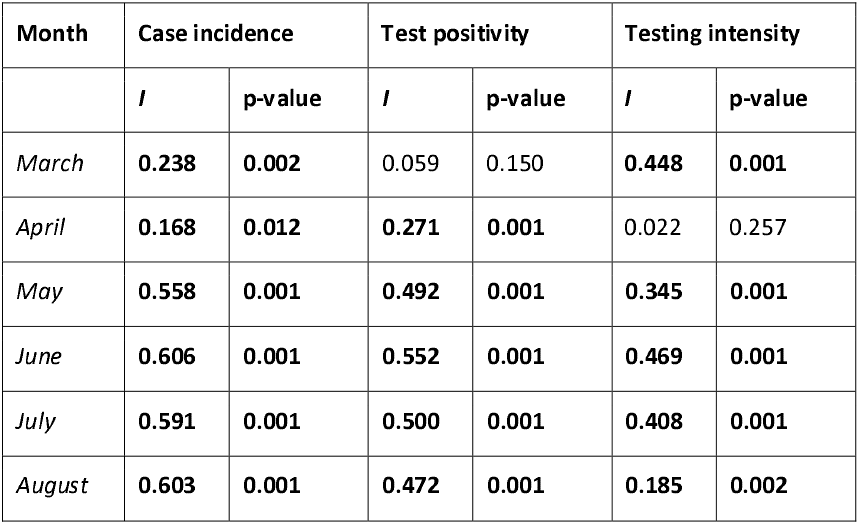
Global Moran’ s / statistics for reported case incidence, test positivity, and testing intensity for each month of the study period (March – August). The / statistic indicates the degree of spatial clustering whereas the simulated p-value gives an indication of statistical significance. Moran’ s / values roughly range from -1 to 1, with 1 indicating complete spatial clustering (i.e. all areas with high values are neighboring other areas with high values) and -1 indicating complete spatial dispersion (with high value areas always neighboring low value areas).

| Month | Case incidence |  | Test positivity |  | Testing intensity |  |
| --- | --- | --- | --- | --- | --- | --- |
|  | *I* | p-value | *I* | p-value | *I* | p-value |
| *March* | **0.238** | **0.002** | 0.059 | 0.150 | **0.448** | **0.001** |
| *April* | **0.168** | **0.012** | **0.271** | **0.001** | 0.022 | 0.257 |
| *May* | **0.558** | **0.001** | **0.492** | **0.001** | **0.345** | **0.001** |
| *June* | **0.606** | **0.001** | **0.552** | **0.001** | **0.469** | **0.001** |
| *July* | **0.591** | **0.001** | **0.500** | **0.001** | **0.408** | **0.001** |
| *August* | **0.603** | **0.001** | **0.472** | **0.001** | **0.185** | **0.002** |

**Figure 1:**
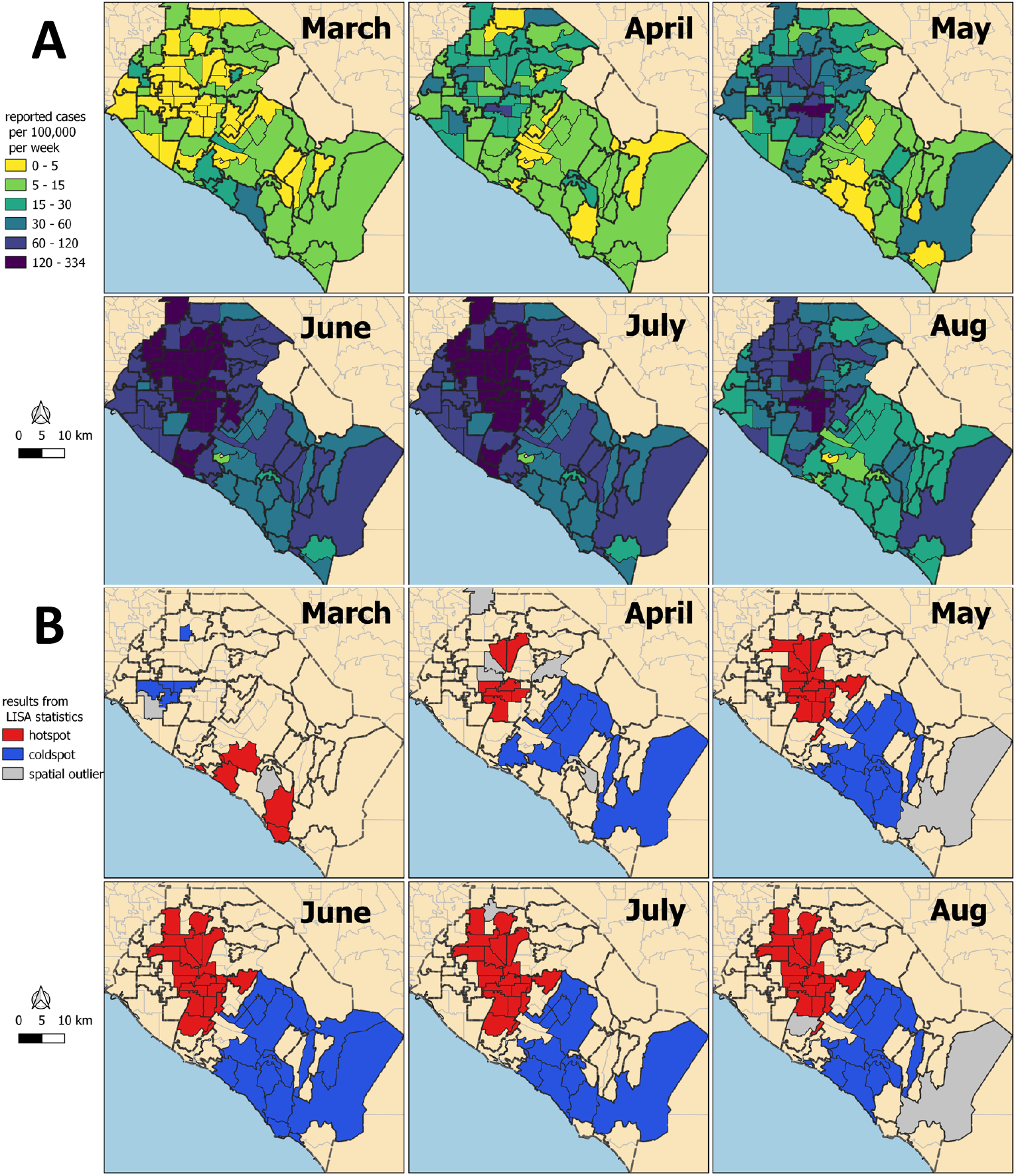
A.) Reported case incidence of COVID-19 in Orange County, California by month. B.) Results from tests of statistical clustering (LISA statistics). Case incidence is calculated as the number of cases per 100,000 people per week.

**Figure 2:**
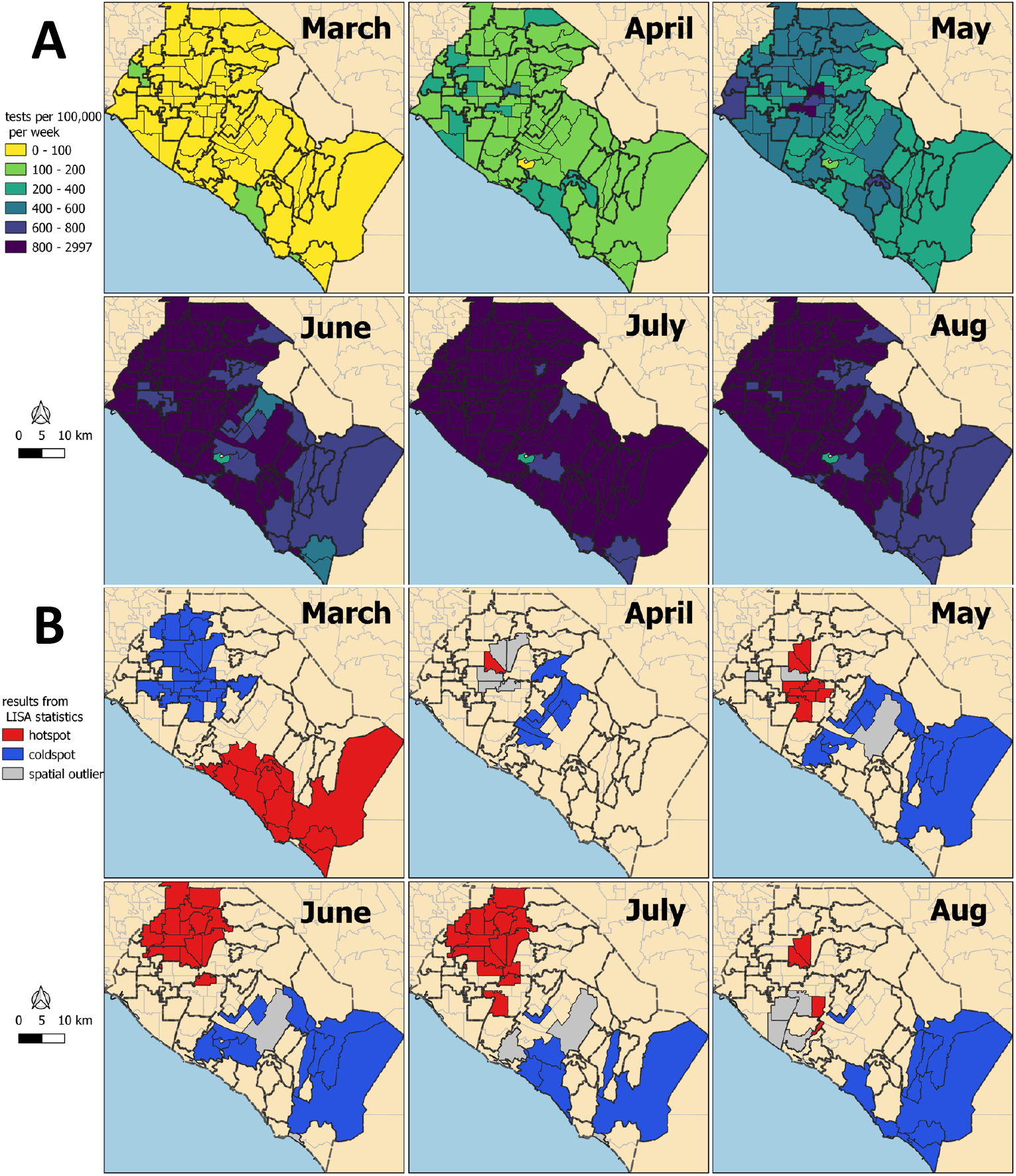
A.) Test intensity in Orange County by month. B.) Results from tests of statistical clustering (LISA statistics). Test intensity is calculated as the number of tests per 100,000 people per week at the zip code level.

Clustering of both reported cases and test positivity increased in magnitude in May (**Table 1** and **Supplementary Figures 3 and 5**). While clustering in test intensity (**Table 1**; **Supplemental Figure 4**) was high in March, it decreased in May as access to testing spread throughout much of the county. Clustering in testing intensity increased again in June and July (centered on the hotspots in the north-central part of the county, evident in **Figures 1 and 3**). By April, case incidence, testing intensity, and test positivity had all shifted to a growing cluster in the north-central part of the county. Zip code level seropositivity also revealed a cluster in the north-central part of OC (**Figure 4**), especially in the city of Santa Ana (**Supplementary Figure 1**).

**Figure 3:**
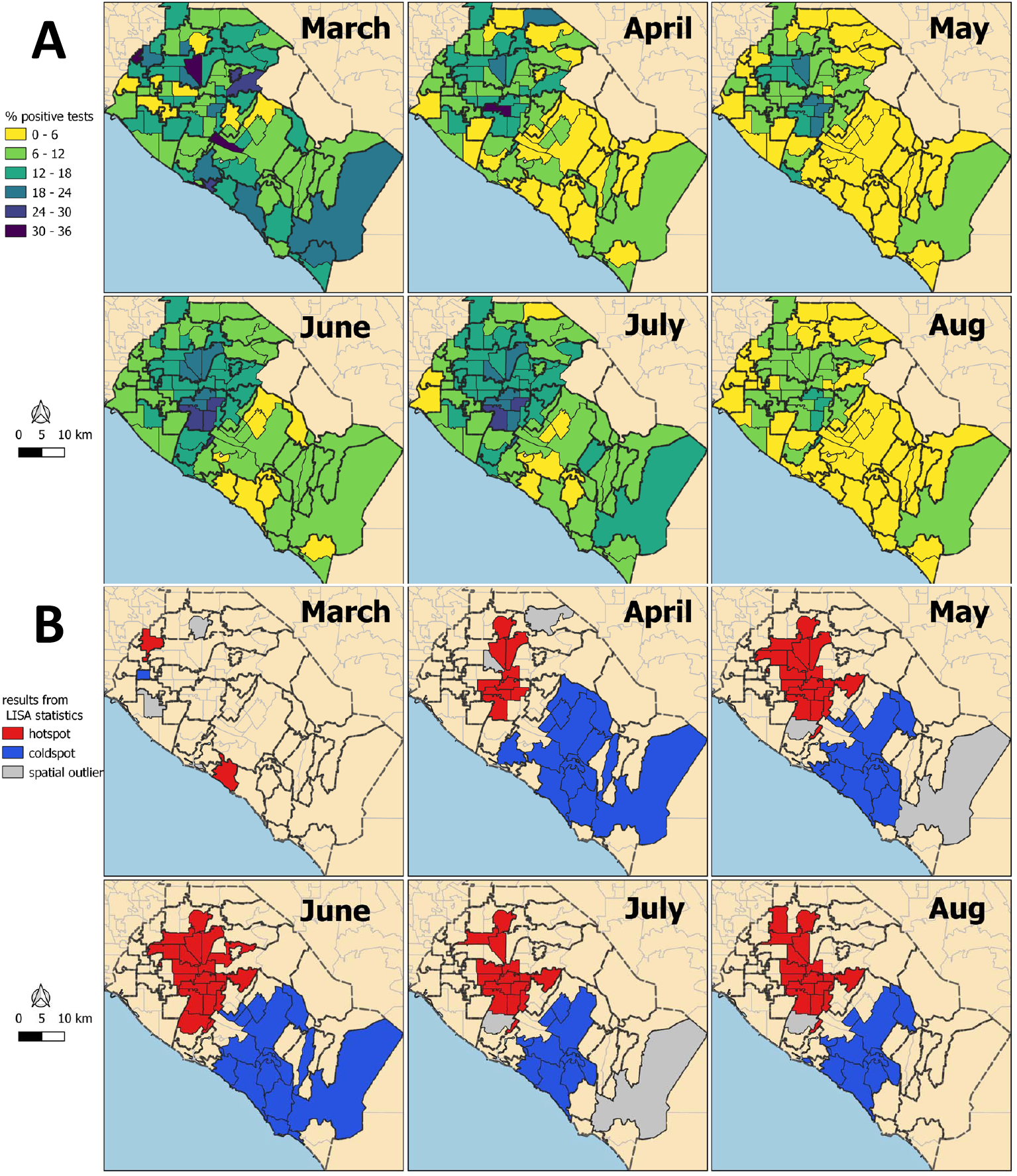
A.) Test positivity (% of tests positive for SARS-CoV-2) at the zip code level in Orange County by month. B.) Results from tests of statistical clustering (LISA statistics).

**Figure 4:**
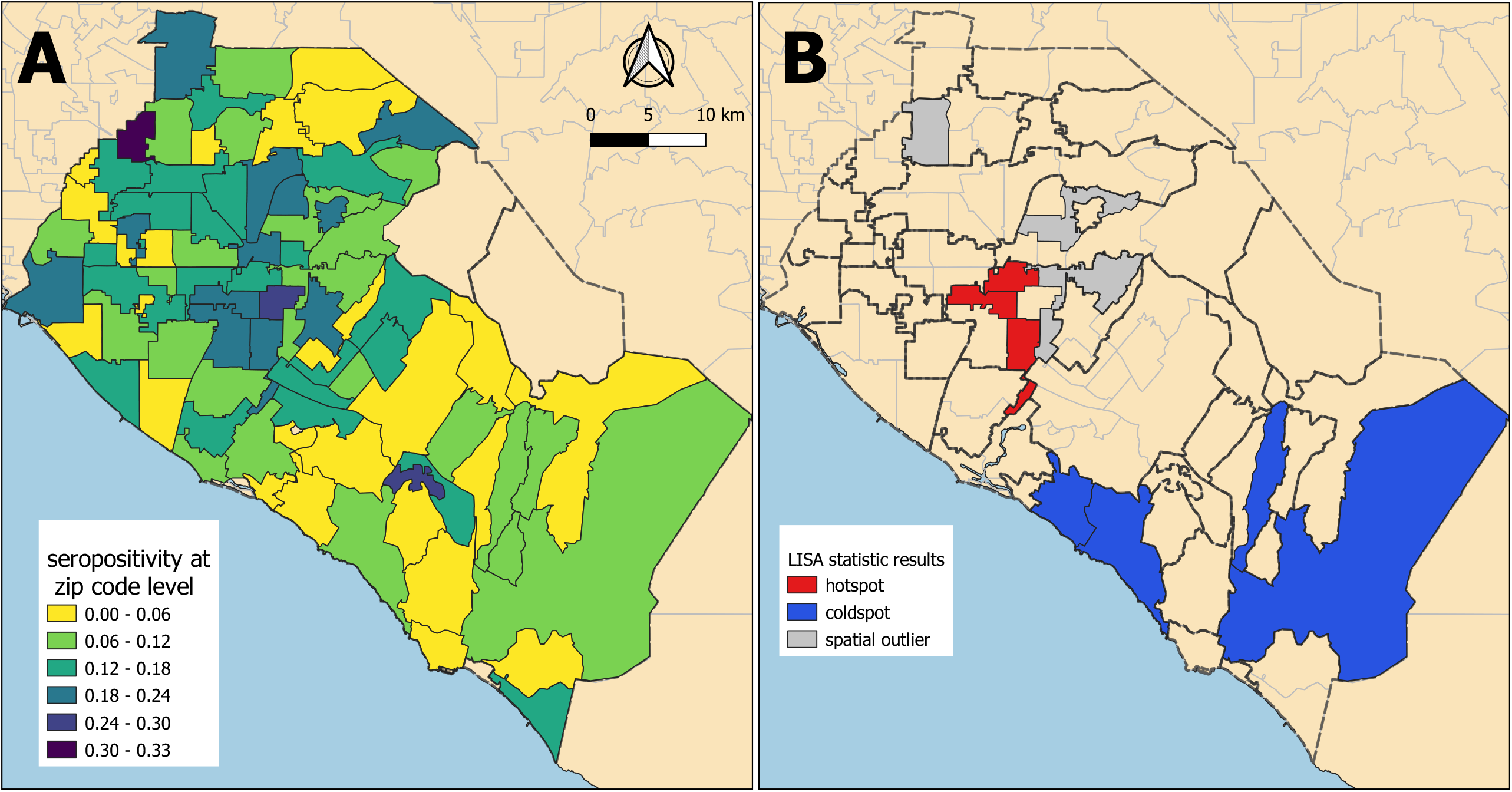
A.) Seropositivity to SARS-CoV-2 at the zip code level, B.) Results of LISA statistics for seropositivity data.

### Results from GAM Logistic Regressions

#### Factors associated with testing positive for SARS-CoV-2 infection

Age was a strong predictor of testing positive. Individuals in the 10-14 and 15-19 age groups had the highest odds of testing positive (both with approximately 2.2 times the odds of testing positive in comparison to the 0 – 4 age group (**Table 2** and **Figure 5**)). Males had 1.2 times the odds of testing positive (95% CI: 1.18 – 1.23). Individuals who identified as Hispanic or Latino had 1.7 times the odds of testing positive (CI: 1.63 – 1.79) when compared to whites, while Asian (AOR: 0.52; CI: 0.49 – 0.55), Black (AOR: 0.58; CI: 0.52 – 0.66), and Pacific Islander (AOR: 0.35; CI: 0.29 – 0.42) individuals had lower odds of testing positive. A large proportion of individuals did not have attributable race or ethnicity data in the records (63 % of all records through August 16).

**Table 2:** Generalized additive logistic regression results for odds of testing positive for SARS-CoV-2 in Orange County. This table excludes the coefficients for median income and time due to the interaction between median income and time. A random intercept was included for zip code.

|  | Counts |  | Adjusted Odds Ratio *<br>with (95% CI†) |
| --- | --- | --- | --- |
|  | SARS-CoV-2+ | Total |  |
| Age |  |  |  |
| 0-4 | 480 (1.3%) | 4813 (1.51%) | Reference |
| 5-9 | 487 (1.32%) | 3841 (1.21%) | 1.60 (1.4, 1.84) |
| 10-14 | 840 (2.28%) | 5037 (1.58%) | 2.23 (1.97, 2.53) |
| 15-19 | 2075 (5.64%) | 13730 (4.31%) | 2.24 (2.01, 2.5) |
| 20-24 | 4529 (12.3%) | 31910 (10.02%) | 1.94 (1.75, 2.15) |
| 25-29 | 4537 (12.32%) | 34997 (10.99%) | 1.64 (1.48, 1.82) |
| 30-34 | 3691 (10.03%) | 30112 (9.45%) | 1.54 (1.39, 1.71) |
| 35-39 | 3220 (8.75%) | 26010 (8.17%) | 1.60 (1.44, 1.77) |
| 40-49 | 5809 (15.78%) | 45255 (14.21%) | 1.68 (1.52, 1.86) |
| 50-59 | 5639 (15.32%) | 48937 (15.37%) | 1.49 (1.35, 1.65) |
| 60-69 | 2964 (8.05%) | 36408 (11.43%) | 1.01 (0.91, 1.13) |
| 70-79 | 1408 (3.82%) | 22204 (6.97%) | 0.76 (0.68, 0.85) |
| 80+ | 1137 (3.09%) | 15238 (4.78%) | 0.79 (0.71, 0.89) |
| Gender |  |  |  |
| Female | 18752 (50.93%) | 174949 (54.93%) | Reference |
| Male | 18064 (49.07%) | 143543 (45.07%) | 1.21 (1.18, 1.23) |
| Race/ethnicity |  |  |  |
| White | 11326 (30.76%) | 62200 (19.53%) | Reference |
| Asian | 1438 (3.91%) | 13764 (4.32%) | 0.52 (0.49, 0.55) |
| Black | 280 (0.76%) | 2055 (0.65%) | 0.58 (0.52, 0.66) |
| Hispanic | 3305 (8.98%) | 9044 (2.84%) | 1.71 (1.63, 1.79) |
| Native American | 48 (0.13%) | 307 (0.1%) | 0.76 (0.56, 1.03) |
| Pacific Islander | 110 (0.3%) | 1583 (0.5%) | 0.35 (0.29, 0.42) |
| Other | 3768 (10.23%) | 29467 (9.25%) | 0.43 (0.41, 0.45) |
| Unknown | 16541 (44.93%) | 200072 (62.82%) | 0.33 (0.32, 0.34) |
| % with College Degree‡ |  |  |  |
| 1st Quartile | 20243 (54.98%) | 121454 (38.13%) | Reference |
| 2nd Quartile | 9284 (25.22%) | 88178 (27.69%) | 0.77 (0.66, 0.9) |
| 3rd Quartile | 4497 (12.21%) | 64915 (20.38%) | 0.59 (0.49, 0.71) |
| 4th Quartile | 2792 (7.58%) | 43945 (13.8%) | 0.61 (0.49, 0.76) |

| % with Insurance |  |  |  |
| --- | --- | --- | --- |
| 1st Quartile | 19270 (52.34%) | 112811 (35.42%) | Reference |
| 2nd Quartile | 10249 (27.84%) | 94061 (29.53%) | 0.74 (0.65, 0.85) |
| 3rd Quartile | 3774 (10.25%) | 53358 (16.75%) | 0.57 (0.47, 0.69) |
| 4th Quartile | 3523 (9.57%) | 58262 (18.29%) | 0.51 (0.41, 0.63) |
| Population Density (1000ppl/km<sup>2</sup>) |  |  | 1.07 (0.99, 1.15) |

\* Adjusted for all covariates listed plus zip code estimated median income and time of test in days. Model intercept represents odds of a white female in the 0 to 4 age group in a zip code in the first quartile of college degree and insured with the average population density. The odds of this individual testing positive for COVID-19 is estimated to be 0.19 (0.16, 0.22)
† 95% Confidence Interval
‡ Estimated: percent of people with a bachelor's degree, percent of people with medical insurance, and population density in an individual's zip code

**Figure 5:**
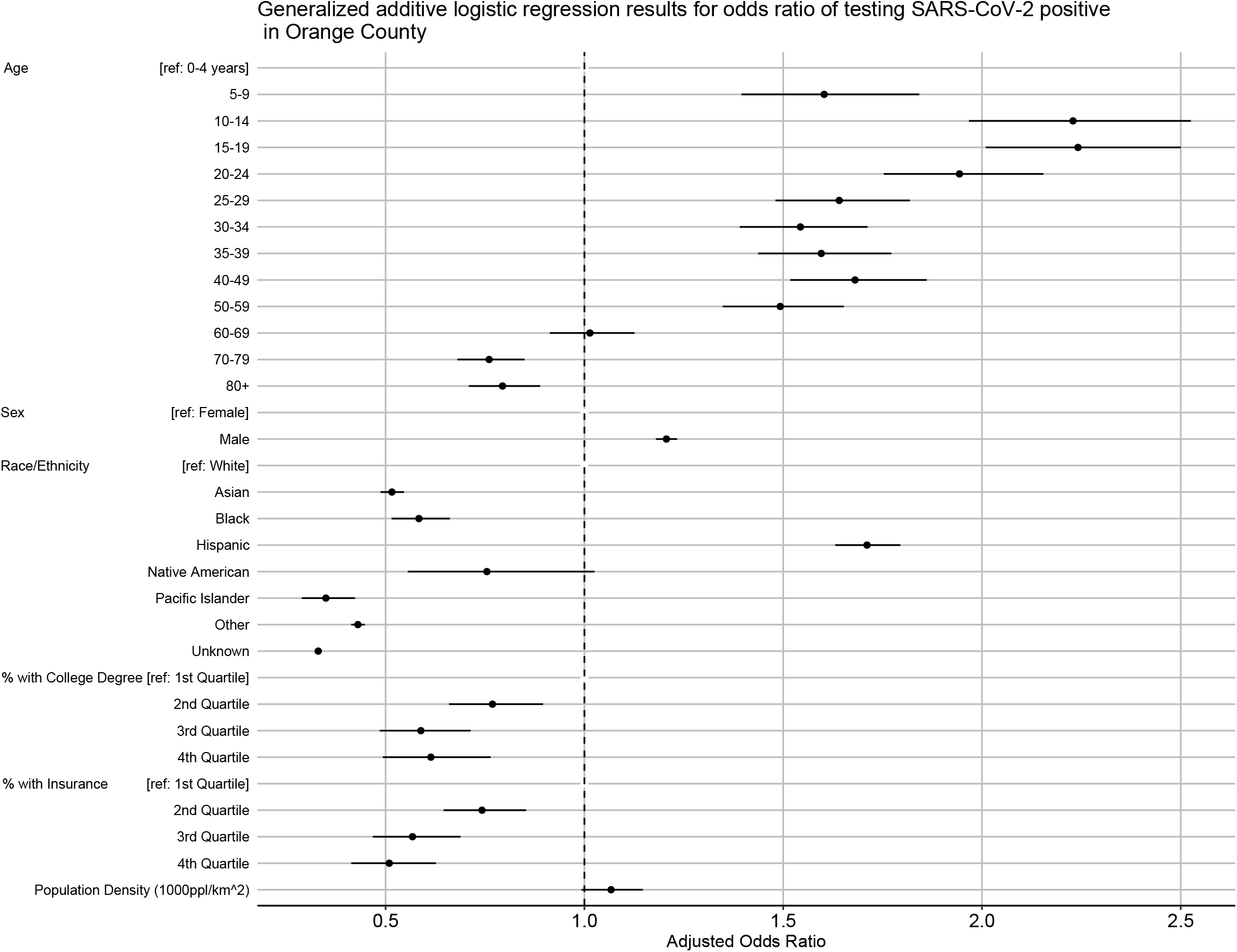
Model adjusted odds ratios and confidence intervals from the logistic regression for odds of testing positive (Table 1)

Zip code level population density was not a significant predictor of testing positive (**Table 2, Figure 5**). However, education (percentage of adults with at least a bachelor’ s degree), insurance coverage (percentage of adults who had insurance in the previous 5 years), and median household income were all statistically significant predictors of testing positive. For example, individuals who lived in zip codes with the highest education levels had 39% decreased odds of testing positive (AOR for the fourth quartile: 0.61; CI: 0.49 – 0.76). In addition, the interaction between zip code level median household income (**Supplementary Figure 2**) indicates that individuals from wealthier zip codes had increased risk of testing positive at the beginning of the OC epidemic. However, this pattern quickly shifted, with individuals from lower income areas showing the highest odds of testing positive in subsequent months.

#### Factors associated with COVID-19 associated mortality

For each increase in 10 years of age there was an associated 2.5 fold increase in the odds of mortality (AOR: 2.56, CI: 2.46 – 2.67; **Table 3, Figure 6**). Infected males were almost twice as likely to die from COVID-19 when compared to females (AOR: 1.97; CI: 1.71 – 2.26). While Asian individuals were less likely to test positive for SARS-CoV-2 infection (**Table 2**), those who did test positive had higher odds of mortality. Compared to whites, Asians had 31% increased odds of dying from COVID-19 (AOR: 1.31; CI: 1.08 – 1.59).

**Table 3.** Logistic regression results for odds of dying among those who tested positive for SARS-CoV-2 in Orange County. A random intercept was included for zip code.

|  | Counts |  | Adjusted Odds Ratio with (95% CI) |
| --- | --- | --- | --- |
|  | COVID-19 Deaths | Total |  |
| Age (decades) |  |  | 2.57 (2.46, 2.68) |
| Gender |  |  |  |
| Female | 542 (43.43%) | 21852 (51.16%) | Reference |
| Male | 706 (56.57%) | 20858 (48.84%) | 2.00 (1.74, 2.29) |
| Race/ethnicity |  |  |  |
| White | 832 (66.67%) | 13894 (32.53%) | Reference |
| Asian | 227 (18.19%) | 2028 (4.75%) | 1.31 (1.08, 1.59) |
| Black | 19 (1.52%) | 347 (0.81%) | 0.95 (0.56, 1.62) |
| Hispanic | 127 (10.18%) | 3917 (9.17%) | 0.94 (0.75, 1.17) |
| Native American | 3 (0.24%) | 60 (0.14%) | 0.71 (0.19, 2.63) |
| Pacific Islander | 6 (0.48%) | 140 (0.33%) | 1.00 (0.41, 2.42) |
| Unknown | 34 (2.72%) | 22324 (52.27%) | 0.03 (0.02, 0.05) |
| % with College Degree |  |  |  |
| 1st Quartile | 768 (61.54%) | 23401 (54.79%) | Reference |
| 2nd Quartile | 259 (20.75%) | 10711 (25.08%) | 0.73 (0.53, 1.01) |
| 3rd Quartile | 178 (14.26%) | 5336 (12.49%) | 0.78 (0.5, 1.21) |
| 4th Quartile | 43 (3.45%) | 3262 (7.64%) | 0.51 (0.27, 0.94) |
| % with Insurance |  |  |  |
| 1st Quartile | 675 (54.09%) | 22170 (51.91%) | Reference |
| 2nd Quartile | 366 (29.33%) | 11997 (28.09%) | 0.83 (0.62, 1.11) |
| 3rd Quartile | 124 (9.94%) | 4403 (10.31%) | 1.11 (0.7, 1.76) |
| 4th Quartile | 83 (6.65%) | 4140 (9.69%) | 0.58 (0.34, 0.99) |
| Population Density (1000ppl/km<sup>2</sup>) |  |  | 1.02 (0.88, 1.2) |
| Median Income (std. dev.) |  |  | 0.96 (0.76, 1.21) |
| Time (std. dev.) |  |  | 1.05 (0.93, 1.19) |
| COVID ICU patients (std. dev.) |  |  | 1.06 (0.97, 1.16) |

\* Model intercept represents odds of death for a white female diagnosed with SARS-CoV-2 in the 0 to 4 age group in a zip code in the first quartile of college degree and insured with the average population density in Orange County. The odds of this individual testing dying is estimated to be 0 (0,0)
† 95% Confidence Interval
‡ Estimated percent of people with a bachelor's degree and with medical insurance in an individual's zip code
§ Percent of hospital beds not being used by COVID-19 patients in Orange County

**Figure 6:**
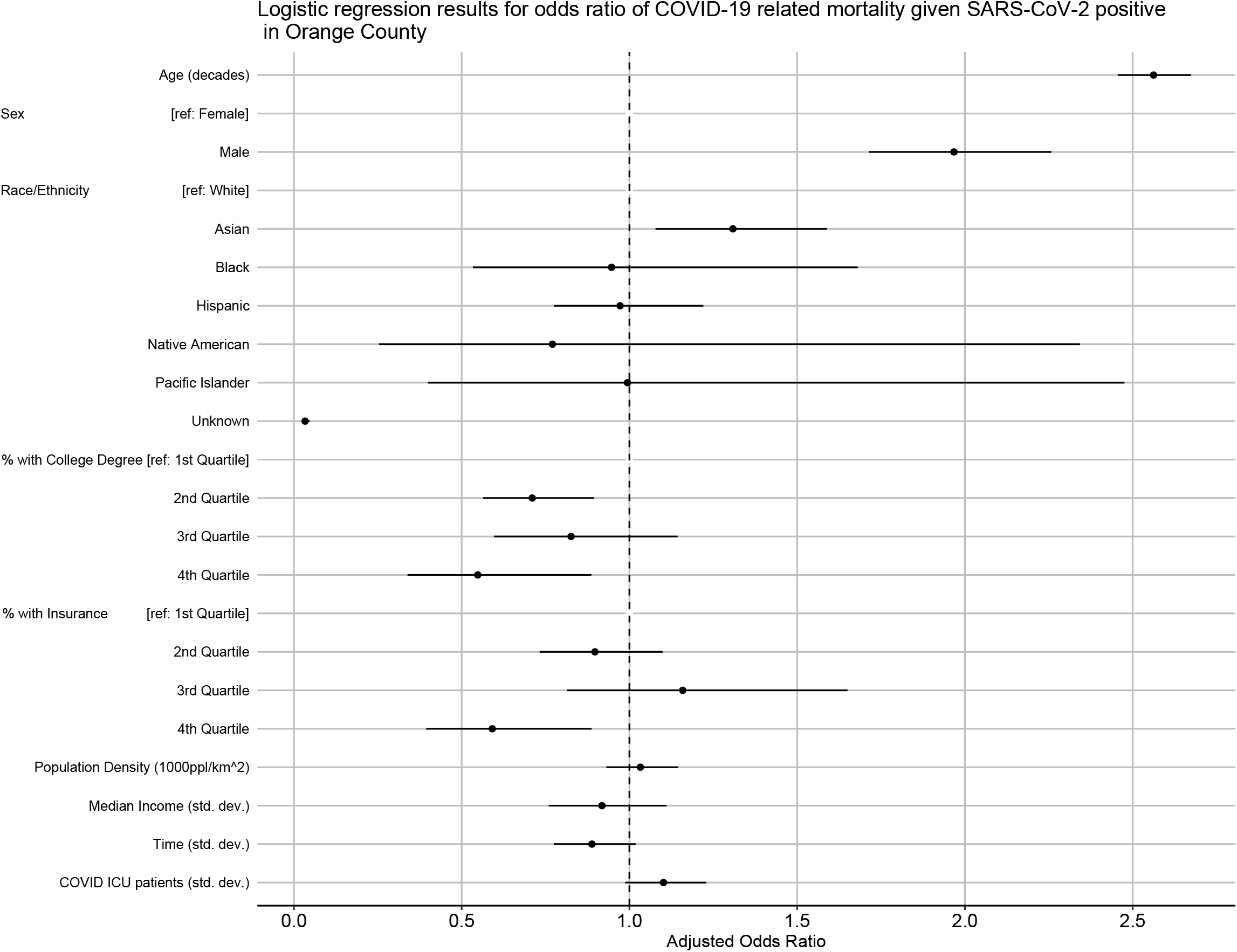
Model adjusted odds ratios and confidence intervals from the logistic regression for the odds of dying from COVID-19

Living in zip codes with high education levels and insurance coverage was also predictive of mortality outcomes (**Table 3, Figure 6**). Individuals who tested positive for COVID-19 and lived in zip codes with the highest levels of educational attainment had 45% lower odds of dying (AOR for the fourth quartile: 0.55; CI: 0.34 – 0.89). Those who lived in zip codes with the highest levels of insurance coverage had 41% lower odds of dying. Zip code level population density and median household income were not significant predictors of mortality after accounting for the other risk factors.

There was no significant change in risk of COVID-19 mortality over this time period. The number of COVID-19 patients in ICU was also not predictive of mortality.

#### Factors associated with SARS-CoV-2 seropositivity

Zip code level cumulative incidence was a significant predictor of individual-level seropositivity in the absence of other zip code level predictors (**Supplementary Table 2**). Every increase in 10% of the zip code cumulative incidence resulted in approximately a 50% increase in the odds that an individual would be seropositive.

Zip code level cumulative incidence was no longer a statistically significant predictor of seropositivity when other zip code level predictors were added to the model (**Table 4, Figure 7**).

**Table 4.** Logistic regression results for odds ratio of testing sero-positive for SARS-CoV-2 in Orange County.

|  | Counts |  | Adjusted Odds Ratio<br>with (95% CI) |
| --- | --- | --- | --- |
|  | SARS-CoV2+ | Total |  |
| Age |  |  |  |
| 18-24 | 19 (5.43%) | 158 (5.35%) | Reference |
| 25-29 | 31 (8.86%) | 234 (7.92%) | 1.08 (0.57, 2.02) |
| 30-34 | 33 (9.43%) | 275 (9.31%) | 0.96 (0.52, 1.8) |
| 35-39 | 35 (10%) | 328 (11.1%) | 0.84 (0.45, 1.55) |
| 40-49 | 83 (23.71%) | 651 (22.04%) | 1.06 (0.61, 1.84) |
| 50-59 | 82 (23.43%) | 659 (22.31%) | 1.07 (0.62, 1.87) |
| 60-69 | 46 (13.14%) | 418 (14.15%) | 1.00 (0.55, 1.83) |
| 70-79 | 18 (5.14%) | 188 (6.36%) | 0.92 (0.45, 1.89) |
| 80+ | 3 (0.86%) | 43 (1.46%) | 0.63 (0.17, 2.26) |
| Sex |  |  |  |
| Female | 222 (63.43%) | 1668 (56.47%) | Reference |
| Male | 128 (36.57%) | 1286 (43.53%) | 0.75 (0.59, 0.94) |
| Race |  |  |  |
| White | 108 (30.86%) | 1228 (41.57%) | Reference |
| Asian | 47 (13.43%) | 435 (14.73%) | 1.26 (0.86, 1.83) |
| Black | 5 (1.43%) | 42 (1.42%) | 1.29 (0.49, 3.4) |
| Hispanic | 162 (46.29%) | 1010 (34.19%) | 1.55 (1.17, 2.03) |
| Pacific Islander | 3 (0.86%) | 12 (0.41%) | 3.94 (1.07, 14.57) |
| Unknown | 25 (7.14%) | 227 (7.68%) | 1.26 (0.78, 2.02) |
| % with College Degree |  |  |  |
| 1st Quartile | 158 (45.14%) | 937 (31.72%) | Reference |
| 2nd Quartile | 92 (26.29%) | 940 (31.82%) | 0.98 (0.65, 1.48) |
| 3rd Quartile | 59 (16.86%) | 596 (20.18%) | 1.31 (0.74, 2.29) |
| 4th Quartile | 41 (11.71%) | 481 (16.28%) | 1.21 (0.63, 2.3) |
| % with Insurance |  |  |  |
| 1st Quartile | 154 (44%) | 928 (31.42%) | Reference |
| 2nd Quartile | 95 (27.14%) | 869 (29.42%) | 0.97 (0.66, 1.42) |
| 3rd Quartile | 50 (14.29%) | 539 (18.25%) | 1.04 (0.57, 1.88) |
| 4th Quartile | 51 (14.57%) | 618 (20.92%) | 0.94 (0.5, 1.77) |
| Population Density (1000ppl/km^2) |  |  | 1.02 (0.84, 1.24) |
| Medina Income (std. dev.) |  |  | 0.75 (0.57, 0.98) |
| % Zip Code SARS-CoV-2+ |  |  | 1.28 (0.95, 1.73) |

\* Model intercept represents odds of testing sero-positive for SARS-CoV2 for a white female diagnosed with SARS-CoV2 in the 18-24 age group in a zip code in the first quartile of college degree and insured with the average population density, and average percent of SARS-CoV2 positive individuals in Orange County. The odds of this individual testing sero-positive is estimated to be 0.074 (0.031,0.178)
† 95% confidence interval computed with robust standard errors
‡ Native American/Native Alaskan race group not included in analysis due to lack of data, no individual of this race group tested seropositive
§ The estimated percent of people with a bachelor's degree, and similarly the estimated percent of people with medical insurance, in an individual's zip code
¶ Number of individuals who tested positive in individual's zip code reported to OC Public Health Department from March 1st to August 16th, divided by estimated population of zip code

**Figure 7:**
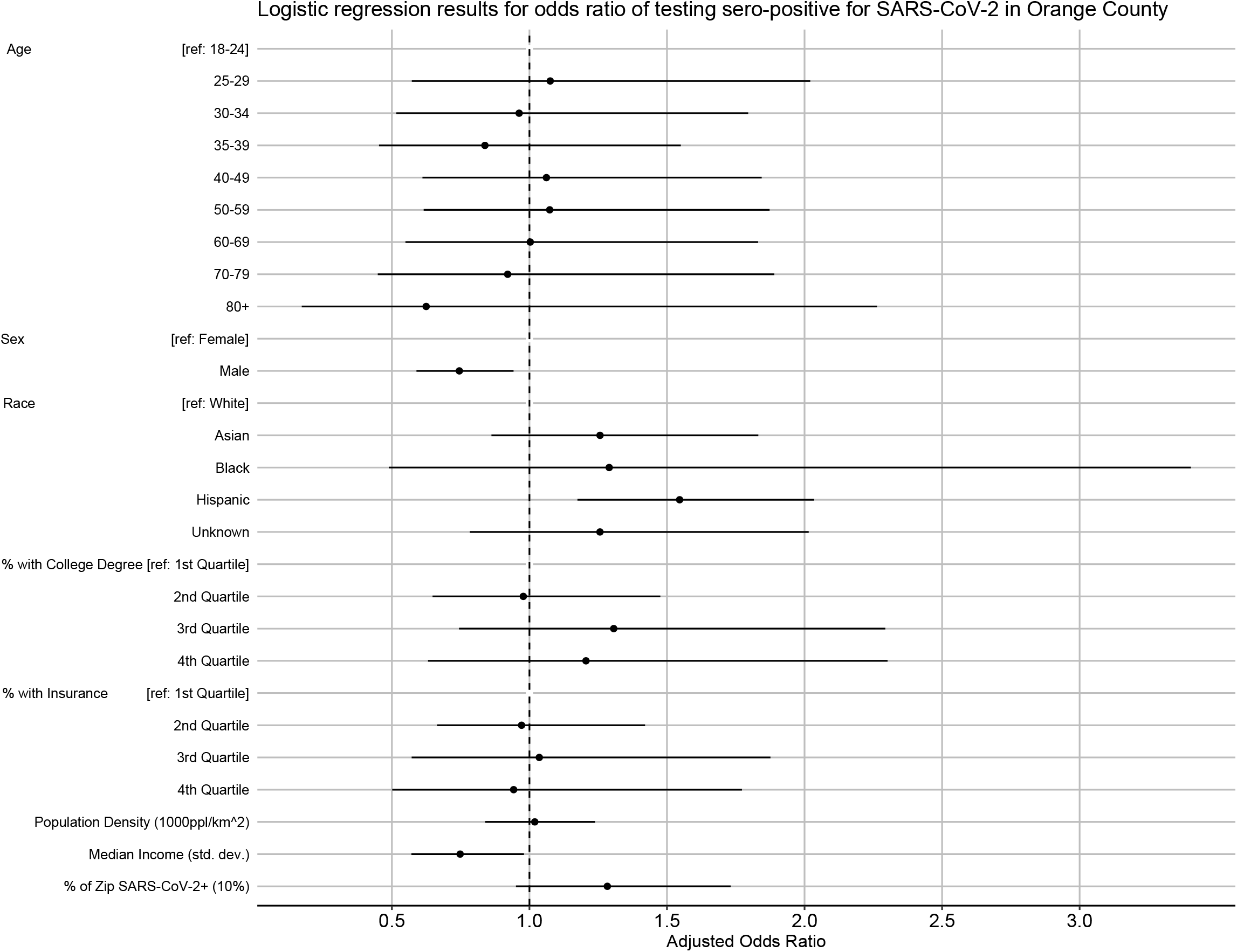
Model adjusted odds ratios and confidence intervals from the logistic regression for the odds of being seropositive for SARS-CoV-2

In the full model (including all zip code level covariates) median household income had a protective effect, with individuals coming from zip codes with higher median household income having lower odds of being seropositive for SARS-CoV-2 antibodies (AOR for every one standard deviation increase: 0.75; CI: 0.57 – 0.98).

We found no difference in age groups with regard to seropositivity. While males were more likely to test positive or to die from SARS-CoV-2 infection, they were less likely than females to be seropositive (AOR: 0.75; CI: 0.59 – 0.94). Hispanic and Latino individuals had 55% increased odds of being seropositive (AOR: 1.55, CI: 1.17 – 2.03). Pacific Islanders may also have had higher odds of being seropositive, but with small total numbers and broad confidence intervals (AOR: 3.94, CI: 1.07 – 14.57; a total of 3 out of 12 individuals tested were seropositive).

## DISCUSSION

Infectious disease data from passive case detection can be biased in a variety of ways, including the well-documented challenge of uneven access to testing and diagnosis (17). In our analysis of COVID-19 in OC, we used a rich set of complementary data that include those passively collected (reported cases, mortality records) and those from active screening (population-based serological testing). Results indicate that, in the early days of the epidemic in OC, both testing intensity and test positivity concentrated in wealthy and affluent areas along the central coast. After March, however, a large cluster of reported cases formed in lower-income North-Central OC (especially the cities of Santa Ana and Anaheim, **Supplementary Figure 1**), growing in size in May and persisting over time. Testing intensity has spread throughout the county during this same time period.

Consistent with emerging reports, we also found that age and male gender strongly predict testing positive and COVID-19 associated mortality (18). Intriguingly, whereas older age groups and males were more likely to have symptomatic disease, our population-based serological survey found that females were more likely than males to be seropositive. Hispanic and Latino individuals had higher risk of infection and testing positive, even after controlling for several zip code level socio-economic factors. Given the consistency of this racial/ethnic finding between the models for test positivity and seropositivity, the risk of being infected with SARS-CoV-2 rises above and beyond the risks of living in a zip code with high transmission or a zip code with low-income and low levels of educational attainment. Other studies also note an increased risk of testing positive for Hispanics and Latinos (19–21). Our seroprevalence survey indicates that in OC, this finding is not an artifact of passive case detection but instead represents a true greater risk of infection for Hispanics and Latinos.

While Asians were less likely to test positive for COVID-19, they were more likely to die when infected. This disparity is consistent with national data, though its cause is uncertain (22). This pattern may reflect discrepancies in outreach communication to these communities or other socio-economic and cultural factors (23,24) and warrants further detailed investigation.

Social determinants of health, defined as “conditions in which people are born, grow, work, live, age, and the wider set of forces and systems,” play a critical role in the creation of disparities related to morbidity, mortality, and quality of life (25). These social determinants include (among other factors) poverty, wealth, educational quality, neighborhood conditions, childhood experience, and social support. Several speculative explanations have been proposed for these sociodemographic patterns related to COVID-19, including living in dense quarters. In addition, as the state and local shelter in place and social distancing policies were mandated, individuals who are independently wealthy or who work in occupations where working from home is a viable option, were more capable of practicing social (more accurately “physical”) distancing. People from low socioeconomic status (SES) areas, by contrast, may have less ability to practice social distancing. Our analyses show that individuals from zip codes with lower overall educational attainment and insurance coverage were more likely to test positive for and die from COVID-19. The association with median household income was more complex and changed over time with regard to test positivity. However, we also find that individuals from zip codes with lower median household income were also more likely to be seropositive for SARS-CoV-2. These findings underscore the importance of understanding contextual factors surrounding infectious disease outbreaks.

Study limitations include that County-reported testing and mortality data did not include individual-level information on income, education, and insurance. These variables were only available at the zip code level. Zip codes are unlikely to adequately represent important spatial units (e.g., neighborhoods, communities). Our measure of population density may also not accurately capture the importance of housing or household density. Missing data on race/ethnicity (63% of all official test records) and small counts of some race/ethnicity groups may have impacted our findings for groups with low counts in this analysis. Even when race/ethnicity data were available, they were broad categories (i.e. Asian rather than specific Asian ethnicities). We also note, however, that the population-based seroprevalence data on SARS-CoV-2 included detailed individual-level information on socio-demographic covariates, which we exploited for our detailed analyses.

Study strengths include the diversity of OC in terms of socioeconomic and demographic predictors, which provide sufficient power to investigate these factors in our analyses. California was also one of the first states to issue an executive order for residents to stay home, providing data for several months when only essential workers were permitted to work outside the home. Our analyses were able to identify temporal shifts in the demographics of COVID-19 test positivity that likely reflect disparities related to occupation type that are further amplified by household characteristics. Finally, we are able to assess differences in risk of infection and test positivity by comparing our population-level serological survey to routinely collected (passive) data from County statistics.

The reasons for the spatial, socio-demographic, and economic patterns we discovered are likely complex and broadly related to issues of accessing health care and general social determinants of health. The clear disparities in how this disease has manifested in OC point toward the need for approaches that are socio-culturally appropriate and those that have a focus on health equity as a fundamental building block. Measures that focus on the hardest-hit communities may serve as efficient points of intervention for COVID-19.

## Data Availability

Data on reported cases and deaths came from the Orange County Health Care Agency (OCHCA). These data can be requested directly through OCHCA:
Alissa Dratch:
The serological data can be requested directly from the PI or MPI of that project:
Dr Tim Bruckner:
Dr Bernadette Boden-Albala

https://github.com/CatalinaMedina/oc-positivity-plus

## ACKNOWLEDGEMENTS

We acknowledge the actOC research manager (Emily Drum) and logistical expertise of Dr Bruce Albala for setting up and helping run the seroprevalence study. Many UCI students and alumni were involved in collecting blood samples for the serological survey and we gratefully acknowledge their contributions. We are grateful to the Felgner lab members who made the serological survey possible under sometimes trying conditions (especially Rie Nakajima, Aarti Jain, and Rafael Ramiro de Assis). Guiyun Yan and Xiaoming Wang provided space for cold storage. The serological survey was funded through a contract with OCHCA (MA-042-20011978). We also acknowledge conversations with colleagues and community leaders with regard to social determinants of health and health equity, including but not limited to: Sora Tanjasiri, Mary Anne Foo, Brittany Morey, Ahn Ellen, Tricia Nguyen, America Bracho, and many others.

## First author biography

Dr. Parker is an infectious disease epidemiologist with expertise in spatial epidemiology, demography, and biomedical anthropology. He is an assistant professor in public health at the University of California, Irvine and the director of the Global Health Research, Education, and Translation (GHREAT) initiative.

## Notes

### Competing Interest Statement

The authors have declared no competing interest.

### Author Declarations

This analysis includes a retrospective analysis of de-identified, anonymized epidemiological records. The UCI Institutional Review Board (IRB) reviewed the details of this study, provided written confirmation that this study is not considered human subjects research, and that the study has been waived from ethics approval.

### Summary of Updates

Added ORCID number for V. Vieira. The original submission was missing a figure and the figure numbers are therefore out of order. I've updated with the correct figures in this version.

## REFERENCES

1. Dowd JB, Andriano L, Brazel DM, Rotondi V, Block P, Ding X, et al. Demographic science aids in understanding the spread and fatality rates of COVID-19. PNAS [Internet]. 2020 Apr 16 [cited 2020 Apr 23]; Available from: https://www.pnas.org/content/early/2020/04/15/2004911117

2. Dorn A van, Cooney RE, Sabin ML. COVID-19 exacerbating inequalities in the US. The Lancet. 2020 Apr 18;395(10232):1243–4.

3. Yancy CW. COVID-19 and African Americans. JAMA [Internet]. 2020 Apr 15 [cited 2020 Apr 23]; Available from: https://jamanetwork.com/journals/jama/fullarticle/2764789

4. Garg S. Hospitalization Rates and Characteristics of Patients Hospitalized with Laboratory-Confirmed Coronavirus Disease 2019 — COVID-NET, 14 States, March 1–30, 2020. MMWR Morb Mortal Wkly Rep [Internet]. 2020 [cited 2020 Apr 23];69. Available from: https://www.cdc.gov/mmwr/volumes/69/wr/mm6915e3.htm

5. Dondorp AM, Hayat M, Aryal D, Beane A, Schultz MJ. Respiratory Support in Novel Coronavirus Disease (COVID-19) Patients, with a Focus on Resource-Limited Settings. The American Journal of Tropical Medicine and Hygiene [Internet]. 2020 Apr 21 [cited 2020 Apr 23]; Available from: http://www.ajtmh.org/content/journals/10.4269/ajtmh.20-0283

6. Bhatraju PK, Ghassemieh BJ, Nichols M, Kim R, Jerome KR, Nalla AK, et al. Covid-19 in Critically Ill Patients in the Seattle Region — Case Series. New England Journal of Medicine. 2020 Mar 30;0(0):null.

7. Yang X, Yu Y, Xu J, Shu H, Xia J, Liu H, et al. Clinical course and outcomes of critically ill patients with SARS-CoV-2 pneumonia in Wuhan, China: a single-centered, retrospective, observational study. The Lancet Respiratory Medicine [Internet]. 2020 Feb 24 [cited 2020 Apr 23];0(0). Available from: https://www.thelancet.com/journals/lanres/article/PIIS2213-2600(20)30079-5/abstract

8. Grasselli G, Pesenti A, Cecconi M. Critical Care Utilization for the COVID-19 Outbreak in Lombardy, Italy: Early Experience and Forecast During an Emergency Response. JAMA [Internet]. 2020 Mar 13 [cited 2020 Apr 23]; Available from: https://jamanetwork.com/journals/jama/fullarticle/2763188

9. Maxmen A. Thousands of coronavirus tests are going unused in US labs. Nature. 2020 Apr 9;580(7803):312–3.

10. Orange County Public Health Order [Internet]. 2020 Mar [cited 2020 Dec 18]. Available from: https://cms.ocgov.com/civicax/filebank/blobdload.aspx?BlobID=114421

11. California S of. Essential workforce [Internet]. [cited 2020 Dec 18]. Available from: https://covid19.ca.gov/essential-workforce/

12. Together OCH. Orange County’ s Healthier Together⍰:: Indicators⍰:: OC Dashboard [Internet]. [cited 2020 Dec 18]. Available from: http://www.ochealthiertogether.org/indicators/index/dashboard?alias=ocdashboard&localeId=267&page=2&card=1

13. Transforming Orange County [Internet]. Transforming Orange County. [cited 2020 Dec 18]. Available from: https://transformingoc.advancingjustice-oc.org

14. Bruckner TA, Parker DM, Bartell SM, Vieira VM, Khan S, Noymer A, et al. Estimated Seroprevalence of SARS-CoV-2 Antibodies Among Adults in Orange County, California. medRxiv. 2020 Oct 12;2020.10.07.20208660.

15. de Assis RR, Jain A, Nakajima R, Jasinskas A, Felgner J, Obiero JM, et al. Analysis of SARS-CoV-2 Antibodies in COVID-19 Convalescent Blood using a Coronavirus Antigen Microarray. bioRxiv [Internet]. 2020 May 8 [cited 2021 Jan 8]; Available from: https://www.ncbi.nlm.nih.gov/pmc/articles/PMC7217240/

16. Anselin L. Local Indicators of Spatial Association. Geographical Analysis. 1995;27:93–115.

17. Zhou G, Afrane YA, Malla S, Githeko AK, Yan G. Active case surveillance, passive case surveillance and asymptomatic malaria parasite screening illustrate different age distribution, spatial clustering and seasonality in western Kenya. Malaria Journal. 2015 Jan 28;14(1):41.

18. Mi J, Zhong W, Huang C, Zhang W, Tan L, Ding L. Gender, age and comorbidities as the main prognostic factors in patients with COVID-19 pneumonia. Am J Transl Res. 2020;12(10):6537–48.

19. Ogedegbe G, Ravenell J, Adhikari S, Butler M, Cook T, Francois F, et al. Assessment of Racial/Ethnic Disparities in Hospitalization and Mortality in Patients With COVID-19 in New York City. JAMA Network Open. 2020 Dec 4;3(12):e2026881.

20. Rubin-Miller L, Alban C, Sep 16 SSP, 2020. COVID-19 Racial Disparities in Testing, Infection, Hospitalization, and Death: Analysis of Epic Patient Data [Internet]. KFF. 2020 [cited 2021 Jan 8]. Available from: https://www.kff.org/coronavirus-covid-19/issue-brief/covid-19-racial-disparities-testing-infection-hospitalization-death-analysis-epic-patient-data/

21. Webb Hooper M, Nápoles AM, Pérez-Stable EJ. COVID-19 and Racial/Ethnic Disparities.JAMA. 2020 Jun 23;323(24):2466.

22. CDC. Coronavirus Disease 2019 (COVID-19) [Internet]. Centers for Disease Control and Prevention. 2020 [cited 2021 Jan 9]. Available from: https://www.cdc.gov/coronavirus/2019-ncov/covid-data/investigations-discovery/hospitalization-death-by-race-ethnicity.html

23. Gover AR, Harper SB, Langton L. Anti-Asian Hate Crime During the COVID-19 Pandemic: Exploring the Reproduction of Inequality. Am J Crim Justice. 2020 Jul 7;1–21.

24. Ng E. The Pandemic of Hate Is Giving Novel Coronavirus Disease (COVID-19) a Helping Hand. Am J Trop Med Hyg [Internet]. 2020 Apr 20 [cited 2021 Jan 8]; Available from: https://www.ncbi.nlm.nih.gov/pmc/articles/PMC7253093/

25. Social determinants of health [Internet]. [cited 2020 Dec 18]. Available from: https://www.who.int/westernpacific/health-topics/social-determinants-of-health

