## Supplemental Materials for "Epidemiology of the early COVID-19 epidemic in Orange County, California: comparison of predictors of test positivity, mortality, and seropositivity" (from another version of this preprint: version 1, 2021-01-15)

**SUPPLEMENTARY MATERIALS**

**Supplementary Table 1**: Description of covariates (predictors) used in logistic regressions

| **Variable** | **description** | **spatial scale** | **transformation** |
|---|---|---|---|
| Age | age of individual | individual level | either age groups or 10 year units |
| Gender | binary indicator for male or female | individual level | |
| Race/ethnicity | race or ethnicity of individual | individual level | |
| % with College Degree | percentage of adults over age 25 with at least a Bachelor's Degree | zip code level | Aggregated into quartiles for multivariable regressions |
| % with Insurance | percentage of adults over 18 with insurance in the last 5 years | zip code level | Aggregated into quartiles for multivariable regressions |
| Population Density | number of people per square kilometer | zip code level | centered on its mean and standardized by standard deviation |
| Median household income | median household income in zip code | zip code level | centered on its mean and standardized by standard deviation |
| % Zip Code SARS-COV-2+ | cumulative incidence of reported COVID-19 cases in zip code, up to August 16th | zip code level | |
| COVID ICU patients (std. dev.) | total number of patients patient with confirmed or suspected COVID-19 in ICU beds (per day) | county level | centered on its mean and standardized by standard deviation |
| Time | continuous variable for time | | centered on its mean and standardized by standard deviation |

**Supplementary Table 2**. Logistic regression results for odds ratio of testing sero-positive for SARS-CoV2 in Orange County. This regression differs from Table 4 in that it excludes zip code level covariates other than the cumulative incidence.

| | Counts | | Adjusted Odds Ratio* with (95% CI†) |
|---|---|---|---|
| | SARS-CoV2+ | Total | |
| Age | | | |
| 18-24 | 19 (5.43%) | 158 (5.35%) | Reference |
| 25-29 | 31 (8.86%) | 234 (7.92%) | 1.113 (0.59, 2.08) |
| 30-34 | 33 (9.43%) | 275 (9.31%) | 0.98 (0.53, 1.82) |
| 35-39 | 35 (10%) | 328 (11.1%) | 0.836 (0.45, 1.54) |
| 40-49 | 83 (23.71%) | 651 (22.04%) | 1.066 (0.62, 1.84) |
| 50-59 | 82 (23.43%) | 659 (22.31%) | 1.071 (0.62, 1.86) |
| 60-69 | 46 (13.14%) | 418 (14.15%) | 0.99 (0.55, 1.8) |
| 70-79 | 18 (5.14%) | 188 (6.36%) | 0.931 (0.46, 1.9) |
| 80+ | 3 (0.86%) | 43 (1.46%) | 0.676 (0.19, 2.42) |
| Sex | | | |
| Female | 222 (63.43%) | 1668 (56.47%) | Reference |
| Male | 128 (36.57%) | 1286 (43.53%) | 0.731 (0.58, 0.92) |
| Race‡ | | | |
| White | 108 (30.86%) | 1228 (41.57%) | Reference |
| Asian | 47 (13.43%) | 435 (14.73%) | 1.286 (0.89, 1.87) |
| Black | 5 (1.43%) | 42 (1.42%) | 1.309 (0.5, 3.43) |
| Hispanic | 162 (46.29%) | 1010 (34.19%) | 1.578 (1.2, 2.08) |
| Pacific Islander | 3 (0.86%) | 12 (0.41%) | 3.62 (1, 13.16) |
| Unknown | 25 (7.14%) | 227 (7.68%) | 1.254 (0.78, 2.01) |
| % Zip Code SARS-CoV2+§ | | | 1.473 (1.24, 1.74) |

* Model intercept represents odds of testing sero-positive for SARS-CoV2 for a white female diagnosed with SARS-CoV2 in the 18-24 age group in a zip code in the first quartile of college degree and insured with the average population density, and average percent of SARS-CoV2 positive individuals in Orange County. The odds of this individual testing sero-positive is estimated to be 0.074 (0.031,0.178)

† 95% confidence interval computed with robust standard errors

‡ Native American/Native Alaskan race group not included in analysis due to lack of data, no individual of this race group tested seropositive

§ Number of individuals who tested positive in individual's zip code reported to OC Public Health Department from March 1st to August 16th, divided by estimated population of zip code

**Supplementary Figure 1**: **Map of Orange County with cities labelled**. Analyses in this research are done at the zip code level (light grey lines indicate zip code boundaries while dashed dark black lines indicate city boundaries).

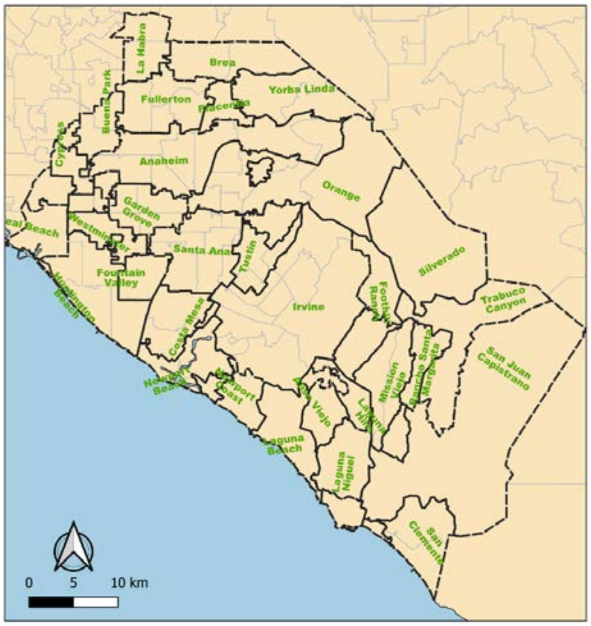


**Supplementary Figure 2**: 3 dimensional plot of the smoothed interaction between zip code level median household income and time as a predictor of testing positive for COVID-19

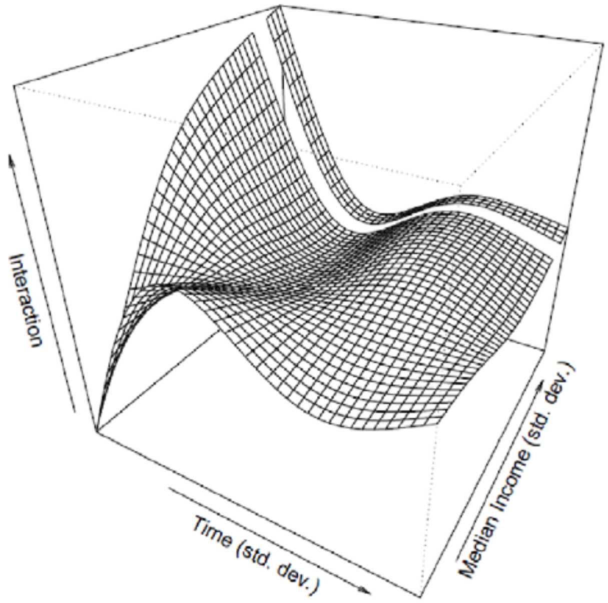

**Supplementary Figure 3**: Spatial correlogram indicating degree of clustering (along y-axis) in reported COVID-19 case incidence by distance (km) away from neighboring zip codes in Orange County, California. Clustering in March is indicated by the black line and subsequent months are indicated by progressively lighter colors. Clustering has been present since the first month (March) but since May the pattern has been stronger and has remained consistent.

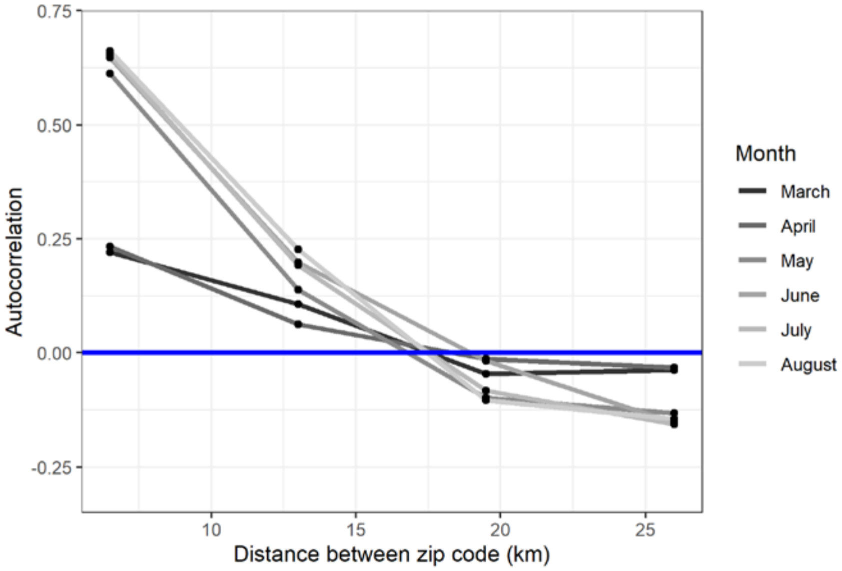


**Supplementary Figure 4**: Spatial correlogram indicating degree of clustering (along y-axis) in COVID-19 testing intensity by distance (km) away from neighboring zip codes in Orange County, California. Clustering in March is indicated by the black line and subsequent months are indicated by progressively lighter colors. Clustering in March was strong and by May was almost non-existent at any spatial extent (May's correlogram falls along the horizontal blue line, with an autocorrelation coefficient near zero). Clustering in testing intensity was again strong in June and July.

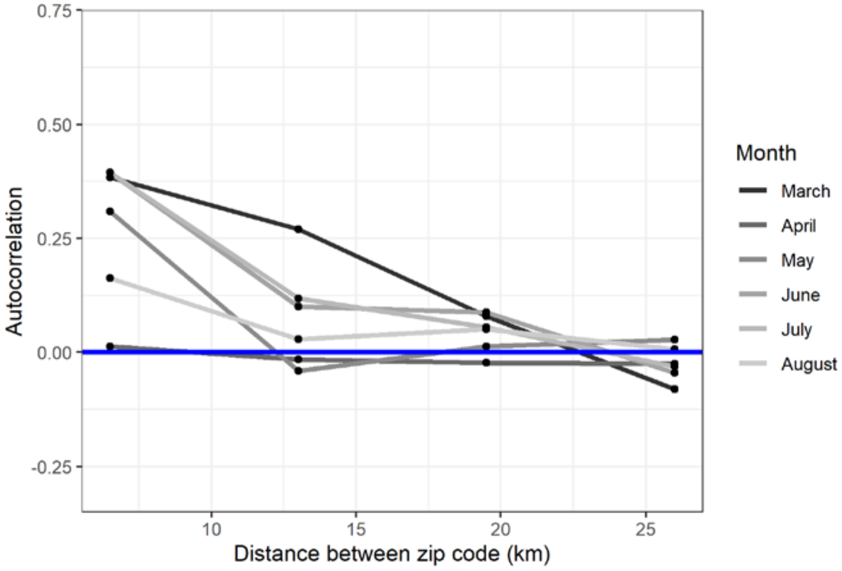

**Supplementary Figure 5**: Spatial correlogram indicating degree of clustering (along y-axis) in COVID-19 test positivity by distance (km) away from neighboring zip codes in Orange County. There was almost no clustering in test positivity in March, at the beginning of the epidemic (the line for March falls near the zero point). Clustering in test positivity was apparent in April, increased in May, and has remained at approximately the same level since.

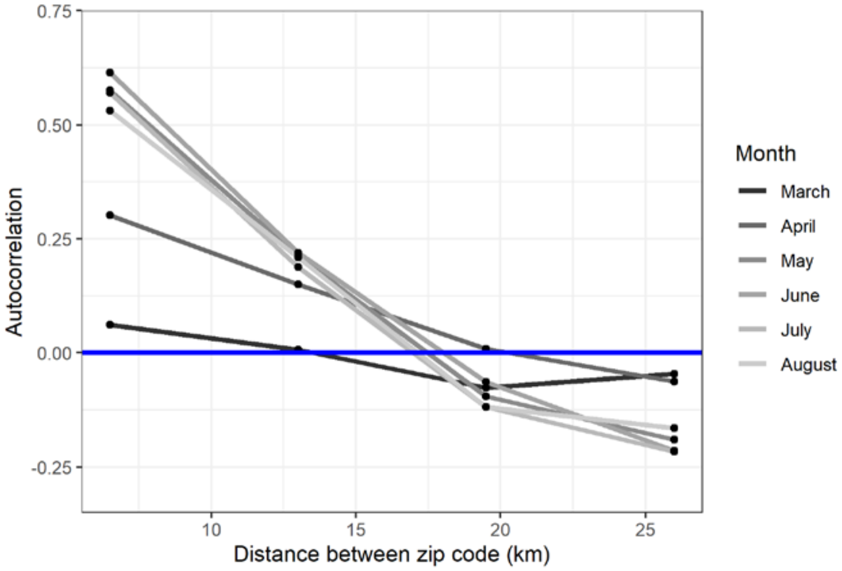

#### Models for testing positive investigation

Let

$OP_i$ be the odds of testing positive for SARS-CoV2 in Orange County.

$\overrightarrow{\beta}_{\text{Age Group}} = (\beta_{\text{Age[5-9]}}, \beta_{\text{Age[10-14]}}, \beta_{\text{Age[15-19]}}, \beta_{\text{Age[20-24]}}, \beta_{\text{Age[25-29]}}, \beta_{\text{Age[30-34]}}, \beta_{\text{Age[35-39]}}$
$\beta_{\text{Age[40-49]}}, \beta_{\text{Age[50-59]}}, \beta_{\text{Age[60-69]}}, \beta_{\text{Age[70-79]}}, \beta_{\text{Age[80+]}})$

$\overrightarrow{\beta}_{\text{Race}} = (\beta_{\text{Asian}}, \beta_{\text{Black}}, \beta_{\text{Hispanic}}, \beta_{\text{NativeAmerican}}, \beta_{\text{PacificIslander}}, \beta_{\text{OtherRace}}, \beta_{\text{Unknown}})$

$\overrightarrow{\beta}_{\text{College}} = (\beta_{\text{\% with College Degree Quartile 2}}, \beta_{\text{\% with College Degree Quartile 3}}, \beta_{\text{\% with College Degree Quartile 4}})$

$\overrightarrow{\beta}_{\text{Insurance}} = (\beta_{\text{\% with Insurance Quartile 2}}, \beta_{\text{\% with Insurance Quartile 3}}, \beta_{\text{\% with Insurance Quartile 4}})$

For clarity the base terms ($\text{BaseTerms}_i$) are defined to be the terms present in all models:

$$\begin{aligned}\text{BaseTerms}_i =& \beta_0 + \overrightarrow{\beta}_{\text{Age Group}} \overrightarrow{\text{Age Group}_i} + \beta_{\text{Sex}}\text{Sex}_i + \overrightarrow{\beta}_{\text{Race}} \overrightarrow{\text{Race}_i} \\ &+ \overrightarrow{\beta}_{\text{College}} \overrightarrow{\text{\% with College Degree Quartile}_i} \\ &+ \overrightarrow{\beta}_{\text{Insurance}} \overrightarrow{\text{\% with Medical Insurance Quartile}_i} \\ &+ \beta_{\text{Population Density}}\text{Population Density}_i + \beta_{\text{Median Income}}\text{Median Income}_i\end{aligned}$$

Model 1:

$$\begin{aligned}\log(OP_i) =& \text{BaseTerms}_i \\ &+ \beta_{\text{Time}}\text{Time}_i,\end{aligned} \tag{1}$$

with a random intercept for zip code.

Model 2:

$$\begin{aligned}\log(O_i) =& \text{BaseTerms}_i \\ &+ \beta_{\text{Time}}\text{Time}_i + \beta_{\text{Time2}}\text{Time}_i^2,\end{aligned} \tag{2}$$

with a random intercept for zip code.

Model 3:

$$\begin{aligned}\log(OP_i) =& \text{BaseTerms}_i \\ &+ \beta_{\text{Time}}\text{Time}_i + \beta_{\text{Time2}}\text{Time}_i^2, \\ &+ \beta_{\text{Interaction1}}\text{Median Income}_i \times \text{Time}_i + \beta_{\text{Interaction2}}\text{Median Income}_i \times \text{Time}_i^2,\end{aligned} \tag{3}$$

with a random intercept for zip code.

Model 4:
Generalized additive model

$$\begin{aligned}\log(OP_i) =& \text{BaseTerms}_i \\ &+ \beta_{\text{Time}} f(\text{Time}_i),\end{aligned} \tag{4}$$

with a random intercept for zip code.

Model 5:
Generalized additive model

$$\begin{aligned}\log(OP_i) =& \text{BaseTerms}_i \\ &+ \beta_{\text{Time}} f(\text{Time}_i) + \beta_{\text{Interaction}}\text{Median Income}_i \times f(\text{Time}_i),\end{aligned} \tag{5}$$

with a random intercept for zip code.

Model 6:
Generalized additive model

$$\begin{aligned}\log(OP_i) =& \text{BaseTerms}_i \\ &+ \beta_{\text{Time}} f(\text{Time}_i) + \beta_{\text{Interaction}}\text{Median Income}_i \times f(\text{Time}_i),\end{aligned} \tag{6}$$

without a random intercept for zip code.

| | Degrees of Freedom | BIC |
|---|---|---|
| Model 1 | 31.00000 | 234798.0 |
| Model 2 | 32.00000 | 233490.8 |
| Model 3 | 34.00000 | 233496.5 |
| Model 4 | 104.07093 | 228996.7 |
| Model 5 | 117.96925 | 228606.7 |
| Model 6 | 51.28528 | 229112.8 |

### Models for mortality investigation

Let
$OM_i$ be the odds of mortality among individuals who tested positive for SARS-CoV2 in Orange County.
$\overrightarrow{\beta}_{\text{Race}} = (\beta_{\text{Asian}}, \beta_{\text{Black}}, \beta_{\text{Hispanic}}, \beta_{\text{Native American}}, \beta_{\text{Pacific Islander}}, \beta_{\text{Other Race}}, \beta_{\text{Unknown}})$
$\overrightarrow{\beta}_{\text{College}} = (\beta_{\text{\% with College Degree Quartile 2}}, \beta_{\text{\% with College Degree Quartile 3}}, \beta_{\text{\% with College Degree Quartile 4}})$
$\overrightarrow{\beta}_{\text{Insurance}} = (\beta_{\text{\% with Insurance Quartile 2}}, \beta_{\text{\% with Insurance Quartile 3}}, \beta_{\text{\% with Insurance Quartile 4}})$.

For clarity the base terms ($\text{BaseTerms}_i$) are defined to be the terms present in all models:

$$\begin{aligned}\text{BaseTerms}_i = \beta_0 &+ \beta_{\text{Age}}\text{Age}_i + \beta_{\text{Sex}}\text{Sex}_i + \overrightarrow{\beta}_{\text{Race}}\overrightarrow{\text{Race}_i} \\ &+ \overrightarrow{\beta}_{\text{College}}\overrightarrow{\text{\% with College Degree Quartile}_i} \\ &+ \overrightarrow{\beta}_{\text{Insurance}}\overrightarrow{\text{\% with Medical Insurance Quartile}_i} \\ &+ \beta_{\text{Population Density}}\text{Population Density}_i + \beta_{\text{Median Income}}\text{Median Income}_i \\ &+ \beta_{\text{Time}}\text{Time}_i \\ &+ \beta_{\text{COVID ICU patients}}\text{COVID ICU patients}_i,\end{aligned}$$

Model 0:

$$\log(OM_i) = \text{BaseTerms}_i \tag{7}$$

without a random intercept for zip code.

Model 1:

$$\log(OM_i) = \text{BaseTerms}_i, \tag{8}$$

with a random intercept for zip code.

| | Degrees of Freedom | BIC |
|---|---|---|
| Model 0 | 19 | 6633.467 |
| Model 1 | 20 | 6628.224 |

#### Models for testing seropositive investigation

Let
$OSP_i$ be the odds of testing positive for COVID-19 in Orange County.
$\overrightarrow{\beta}_{\text{Age Group}} = (\beta_{\text{Age[18-24]}}, \beta_{\text{Age[25-29]}}, \beta_{\text{Age[30-34]}}, \beta_{\text{Age[35-39]}}, \beta_{\text{Age[40-44]}}, \beta_{\text{Age[45-49]}}, \beta_{\text{Age[50-54]}}, \beta_{\text{Age[55-59]}}, \beta_{\text{Age[60-64]}}, \beta_{\text{Age[65-69]}}, \beta_{\text{Age[70-74]}}, \beta_{\text{Age[75-79]}}, \beta_{\text{Age[80-84]}}, \beta_{\text{Age[85+]}})$
$\overrightarrow{\beta}_{\text{Race}} = (\beta_{\text{Asian}}, \beta_{\text{Black}}, \beta_{\text{Hispanic}}, \beta_{\text{PacificIslander}}, \beta_{\text{Unknown}})$
$\overrightarrow{\beta}_{\text{College}} = (\beta_{\text{\% with College Degree Quartile 2}}, \beta_{\text{\% with College Degree Quartile 3}}, \beta_{\text{\% with College Degree Quartile 4}})$
$\overrightarrow{\beta}_{\text{Insurance}} = (\beta_{\text{\% with Insurance Quartile 2}}, \beta_{\text{\% with Insurance Quartile 3}}, \beta_{\text{\% with Insurance Quartile 4}})$

For clarity the base terms ($\text{BaseTerms}_i$) are defined to be the terms present in all models:

$$\begin{aligned}\text{BaseTerms}_i &= \beta_0 + \overrightarrow{\beta}_{\text{Age Group}}\overrightarrow{\text{Age Group}}_i + \beta_{\text{Sex}}\text{Sex}_i + \overrightarrow{\beta}_{\text{Race}}\overrightarrow{\text{Race}}_i \\ &+ \overrightarrow{\beta}_{\text{College}}\overrightarrow{\text{\% with College Degree Quartile}}_i \\ &+ \overrightarrow{\beta}_{\text{Insurance}}\overrightarrow{\text{\% with Medical Insurance Quartile}}_i \\ &+ \beta_{\text{Population Density}}\text{Population Density}_i + \beta_{\text{Median Income}}\text{Median Income}_i,\end{aligned}$$

Model 0:

$$\log(O_i) = \text{BaseTerms}_i, \tag{9}$$

without a random intercept for zip code.

Model 1:

$$\log(O_i) = \text{BaseTerms}_i, \tag{10}$$

with a random intercept for zip code.

Model 2:

$$\begin{aligned}\log(O_i) &= \text{BaseTerms}_i \\ &+ \beta_{\text{Passive detection}}\text{\% of Zip Code SARS-CoV2+}_i,\end{aligned} \tag{11}$$

without a random intercept for zip code.

Model 3:

$$\log(O_i) = \beta_0 + \beta_{\text{Passive detection}}\text{\% of Zip Code SARS-CoV2+}_i, \tag{12}$$

without a random intercept for zip code.

Model 4:

$$\begin{aligned}\log(O_i) &= \beta_0 + \beta_0 + \overrightarrow{\beta}_{\text{Age Group}}\overrightarrow{\text{Age Group}}_i + \beta_{\text{Sex}}\text{Sex}_i + \overrightarrow{\beta}_{\text{Race}}\overrightarrow{\text{Race}}_i \\ &+ \beta_{\text{Passive detection}}\text{\% of Zip Code SARS-CoV2+}_i,\end{aligned} \tag{13}$$

without a random intercept for zip code.

| | Degrees of Freedom | BIC |
|---|---|---|
| Model 0 | 23 | 2268.804 |
| Model 1 | 24 | 2276.795 |
| Model 2 | 24 | 2274.407 |
| Model 3 | 2 | 2129.994 |
| Model 4 | 16 | 2219.442 |
